# Early life factors documented in electronic health records predict recurrent acute otitis media

**DOI:** 10.64898/2026.03.07.26347843

**Authors:** Jillian H. Hurst, Congwen Zhao, Eileen M. Raynor, Janet Lee, Sarah A. Gitomer, Christopher W Woods, Matthew S. Kelly, Michael J. Smith, Benjamin A. Goldstein

## Abstract

**Background and Objectives:** Recurrent acute otitis media (rAOM; defined as ≥3 AOM episodes in 6 months or ≥4 episodes in 12 months) affects 10-15% of children in the United States and is a leading cause of healthcare utilization and antibiotic prescriptions. Prospective identification of children at risk of rAOM could help target interventions and identify new risk factors to guide preventive approaches. We therefore sought to develop predictive models to identify children at risk of rAOM using electronic health records (EHR) data.

**Methods:** We extracted retrospective EHR data for children who were born at Duke University Health System (DUHS) hospitals between January 1, 2014, and June 30, 2022, and who had at least one AOM episode during the study period. We used LASSO to build predictive models for development of rAOM at each episode and identified factors associated with rAOM.

**Results:** We identified 6,566 children who met the study criteria, including 1,634 (24.8%) who met criteria for rAOM. A model using only data available at the first AOM episode had an area under the curve (AUC) of 0.75 (0.73, 0.77) and an Area Under the Precision Recall Curve (AUPRC) of 0.41 (95% CI 0.37, 0.46), indicating moderate discriminative ability. At the time of the first AOM episode, features associated with subsequent rAOM development included age, number of prior antibiotic prescriptions, and diagnosis of gastroesophageal reflux disease (GERD). Further, children who developed rAOM were more likely to experience treatment failure than children who did not meet rAOM criteria across all episodes.

**Conclusions:** Our findings indicate that clinical exposures and patient characteristics documented in the EHR distinguish children who are at risk of developing rAOM. Such models could be deployed within EHR systems to identify children who would benefit from early evaluation by an otolaryngologist and audiologist.

## INTRODUCTION

Acute otitis media (AOM) is one of the most common infectious diseases in early childhood and the leading cause of pediatric medical consultation and antibiotic prescriptions globally.^1–4^ While most children experience only one or two AOM episodes in the first few years of life, an estimated 5-30% of children develop recurrent AOM (rAOM), defined as three or more episodes within a 6-month period or four or more episodes in a year.^5,6^ Recurrent infections have a significant impact on quality of life for children and their families due to frequent healthcare visits, medical costs, and work and school absenteeism.^7–10^ Further, rAOM is accompanied by middle ear effusion, which can impair hearing, potentially resulting in language delays and auditory processing disorders that have lifelong impacts.^11–14^

Despite its high prevalence and negative impacts on children and families, the factors that influence susceptibility to rAOM are not well defined. Known risk factors include household tobacco smoke exposures, male sex, family history of AOM, and daycare attendance, while breastfeeding is a protective factor.^5,15–23^ Prior studies have found inconsistent associations between rAOM and delivery mode, atopic disease, and gastroesophageal reflux disease, potentially due to relatively small cohort sizes and differences in diagnostic criteria.^24–32^ Moreover, rAOM risk is likely to be multifactorial, with risk factors and protective factors changing with age and developmental stage. Given the healthcare burdens associated with this condition, new approaches are needed to identify potentially modifiable risk factors for rAOM.

Electronic health records (EHR) data offer a comprehensive and longitudinal view of a child’s health status, as they capture key data elements and health indicators across multiple healthcare encounters such as regular well child visits. We hypothesized that an analysis of features captured within the EHR from birth through the first few years of life, when children are at greatest risk of AOM, could identify factors that distinguish children at risk of rAOM. Using longitudinal EHR data from over 6000 children with regularly documented care from birth through 4 years, we compared clinical and sociodemographic features between children who met criteria for rAOM and children with one or more documented AOM episodes but who did not meet rAOM criteria. To understand how these factors might relate to rAOM risk, we built interpretable least absolute shrinkage and selection operator (LASSO)-based models to predict which children would go on to meet rAOM criteria at the time of their first, second, third, and fourth AOM episodes.

## METHODS

### Study population

This study was conducted using retrospective EHR data from the Duke University Health System (DUHS). DUHS consists of one tertiary care and two community hospitals, and a network of over 100 primary care and specialty clinics that have utilized a single Epic EHR system since 2014. DUHS is the primary health care provider in Durham County, North Carolina, with an estimated 85% of children in the county receiving care from DUHS.^33^ We abstracted clinical data from Duke’s EHR-based Clinical Research Datamart (CRDM) from January 1, 2014, through June 1, 2022.^34^ We identified children born at DUHS hospitals between January 1, 2014, and June 1, 2018, and obtained data for each child from birth through 48 months of age. To limit the cohort to children who were likely to receive the majority of their care through DUHS, we required that each child have at least four well child visits during the study period, with at least one well child visit in the third or fourth year of life (**Figure 1**). We used ICD-9/10 codes to exclude children with chronic conditions that could influence their susceptibility to AOM, including children with cleft lip and/or palate, Trisomy 21, or immune deficiency; we additionally excluded children with incomplete birth data (**Figure 1**, **Supplemental Table 1**). Finally, the cohort was limited to children who had at least one AOM episode in the first four years of life.

**Figure 1.**
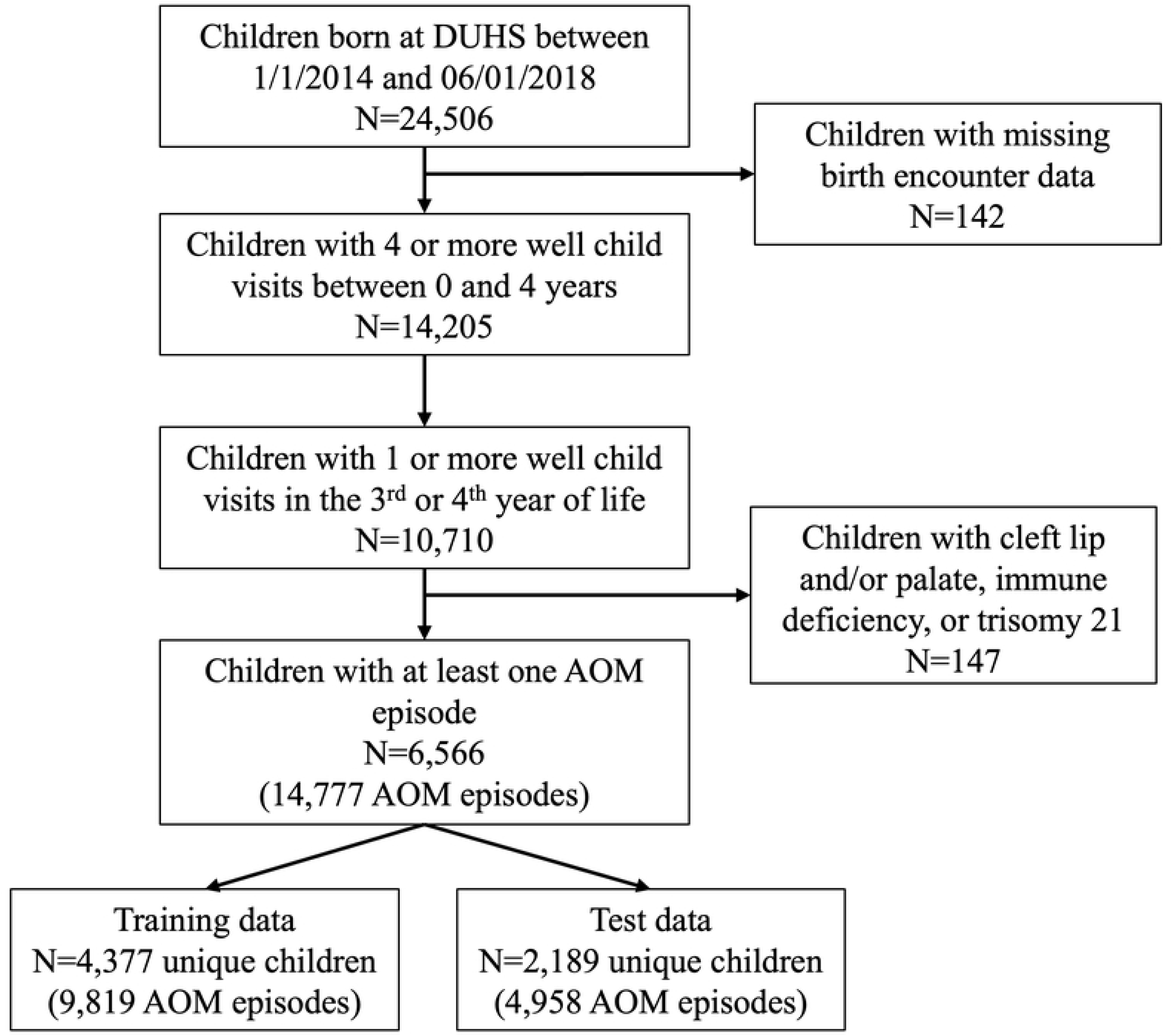
CONSORT diagram for the study cohort. All children included in the cohort were required to have been born at a Duke University-affiliated hospital and to have 4 or more well child visits during the first 4 years of life, including at least 1 visit in their third or fourth year. Children with missing birth data and those who had chronic conditions that could influence their susceptibility to AOM were excluded from analysis.

### Outcome of interest

Our primary outcome of interest was the development of rAOM, defined as three or more AOM episodes within a 6-month period or four or more episodes within a 12-month period.^6^ AOM episodes were identified by ICD-9/10 codes and included encounters in primary care, urgent care, and emergency department visits (**Supplemental Table 1**). Episodes that occurred within a 2-week window were counted as a single episode, as chart review confirmed that the majority of encounters with an AOM diagnosis occurring within this period were for follow-up consultation and/or treatment failure. In these cases, only the first encounter was included in the dataset.

### Data formatting and predictor variables

Our goal was to identify features that predict the likelihood that a child will meet rAOM criteria at the time of each of the first four AOM episodes. We abstracted clinical data from Duke Clinical Research Datamart for each child at birth and throughout the study period, with predictors updated at the time of each AOM episode (**Supplemental Table 1**). Predictor variables at the time of birth included sex, age, gestational age at birth, birth weight, and NICU admission. We additionally extracted maternal information at the time of delivery, including age, delivery mode, ICU admission, and comorbidities and conditions at delivery, including fever, group B streptococcus (GBS) colonization status, chorioamnionitis, gestational diabetes, and tobacco use. We abstracted the following health information for each child over the course of the study period, with information updated at the time of each AOM episode: age, number of prior AOM diagnoses; vaccine doses; antibiotic prescriptions and administrations, including the type of medication, route of administration, and antibiotic spectrum index (ASI); and healthcare utilization history.^35^ AOM episodes were included up to the point at which individuals met criteria for rAOM or the end of the study period.

### Statistical analysis

We first described the demographic and clinical characteristics of children who met criteria for rAOM during the study period compared to those who only had sporadic AOM episodes. Continuous variables were summarized using mean and standard deviation. Categorical variables were presented as numbers and percentages. The standardized mean difference (SMD) was used to measure the magnitude of differences between children who did versus did not go on to meet criteria for rAOM at the time of their first documented AOM episode, with a SMD greater than 0.10 indicating an imbalance between groups. SMDs of 0.2, 0.5, and 0.8 considered to be small, medium, and large differences, respectively.

To develop the predictive models, we divided patients into testing and training sets at a 2:1 ratio. AOM episodes from the same patient were grouped together into either the testing or the training dataset. We fitted LASSO-based models to predict whether a given child would develop rAOM at the time of the first, second, third, and fourth documented AOM episodes. Each model was trained using the data available up to the time of a given episode (e.g., data from birth up to the time of the first episode, data from birth up to the time of the second episode, etc.), resulting in four different episode-specific models. These models were evaluated using data for the corresponding episode within the testing data. We additionally built a model using all available data, regardless of the episode order (the “overall” model), which was evaluated using test data from the first, second, third, and fourth documented AOM episodes.

Based on the prediction of the testing data, we compared the performances of the episode-specific models using three metrics: Area under the receiver operator characteristics (AUROC), area under the precision-recall curve (AUPRC), and calibration slope. AUROC is the probability that a case has a higher risk score than a control and provides a description of the model’s ability to discriminate between cases and non-cases. Area Under the Precision - Recall Curve (AUPRC), which estimates the average positive predictive value (i.e., the fraction of cases labeled as positive that have rAOM) and provides an understanding of how well we can identify a true positive case over random chance. Calibration slope estimates how well a model’s predicted probabilities align with observed outcomes. We used bootstrap resampling at the patient-level to calculate 95% confidence intervals for all metrics. Additionally, we compared the feature importance across each model in order to identify factors associated with rAOM at the time of each AOM episode. All analyses were performed in R 4.1.3, including the *glmnet* package, which was used to fit the LASSO models, and the *ROCR* package, which was used to evaluate model performance.^36–38^

### Ethical Statement

The Duke University Health System Institutional Review Board determined this study protocol to be exempt human subjects research and granted a waiver of informed consent for a retrospective analysis of existing patient data (Pro00112534). Data access began on February 2, 2023. Authors had access to patient identifiers; therefore, all analyses were conducted in a protected analytics environment with the minimum number of identifiers necessary for conduct of the study. All experimental protocols were performed in accordance with relevant guidelines and regulations and approved by the Duke University Health System Internal Review Board.

## RESULTS

### Sociodemographic and clinical characteristics of children with recurrent versus sporadic AOM

A total of 6,566 children were eligible for inclusion, including 1,634 (25%) children who met criteria for rAOM (**Table 1**). We found meaningful differences in race/ethnicity between the two groups (SMD=0.484), with children who identified as non-Hispanic white being more likely to meet rAOM criteria than children of other races and ethnicities. We did not identify any meaningful differences in the prevalence of pre-term birth, NICU admission, receipt of intrapartum antibiotics, or maternal delivery complications (i.e., group B streptococcus (GBS) colonization, maternal fever, chorioamnionitis, or gestational diabetes) between children with and without rAOM. Caesarean delivery was slightly more prevalent among children with rAOM (37% among children with recurrent AOM vs. 31% among children with sporadic AOM, SMD=0.126). Children who met criteria for rAOM had more outpatient healthcare encounters compared to children with sporadic AOM, including well child visits (SMD=0.258) and other outpatient and telehealth encounters (SMD=0.627). Further, children who developed rAOM had their first episode at a younger age (357 days) compared to children with only sporadic AOM episodes (592 days, SMD=0.764). Because children are vaccinated against multiple respiratory pathogens in the first year of life, we specifically evaluated vaccine receipt at the time of first AOM episode. Children who went on to develop rAOM had received fewer pneumococcal conjugate vaccine (PCV) doses (SMD=0.480), fewer *Haemophilus influenzae* type B (Hib) vaccine doses (SMD = 0.240), and fewer seasonal influenza vaccine doses (SMD = 0.484) at the time of their first AOM episode compared to children with sporadic AOM, primarily due to their younger age.

**Table 1.** Patient-level characteristics at first AOM episode.

| Table 1. Patient-level characteristics at first AOM episode |  |  |  |  |
| --- | --- | --- | --- | --- |
| Characteristics and exposures |  | Sporadic AOM | Recurrent AOM | SMD |
|  |  | N = 4,932 | N = 1,634 |  |
| Female infant sex, n (%) |  | 2,388 (48.4%) | 695 (42.5%) | 0.118 |
| Infant race/ethnicity, n (%) |  |  |  | 0.484 |
|  | Hispanic | 879 (17.8%) | 144 (8.8%) |  |
|  | Non-Hispanic white | 1,760 (35.7%) | 947 (58.0%) |  |
|  | Non-Hispanic Black | 1,623 (32.9%) | 342 (20.9%) |  |
|  | Other race/ethnicity | 670 (13.6%) | 201 (12.3%) |  |
| Birth season |  |  |  |  |
|  | Winter (December, January, February) | 1,207 (24.5%) | 389 (23.8%) | 0.052 |
|  | Spring (March, April, May) | 1,311 (26.6%) | 453 (27.7%) |  |
|  | Summer (June, July, August) | 1,235 (25.0%) | 380 (23.3%) |  |
|  | Fall (September, October, November) | 1,179 (23.9%) | 412 (25.2%) |  |
| Pre-term birth (<37 weeks completed), n (%) |  | 497 (10.1%) | 157 (9.6%) | 0.016 |
| Infant admitted to NICU, n (%) |  | 443 (9.0%) | 117 (7.2%) | 0.067 |
| Birthweight, g, mean (sd) |  | 3256.9 (619.9) | 3321.4 (626.9) | 0.103 |
| Caesarean delivery, n (%) |  | 1,539 (31.2) | 607 (37.1) | 0.126 |
| Intrapartum antibiotics, n (%) |  | 1,672 (33.9%) | 521 (31.9%) | 0.043 |
| Maternal delivery conditions, n (%) |  |  |  |  |
|  | Group B Strep-positive | 812 (16.5%) | 263 (16.1%) | 0.010 |
|  | Fever | 49 (1.0%) | 13 (0.8%) | 0.021 |
|  | Chorioamnionitis | 297 (6.0%) | 88 (5.4%) | 0.027 |
|  | Gestational diabetes | 551 (11.2%) | 179 (11.0%) | 0.007 |
|  | Tobacco use during pregnancy | 288 (5.8%) | 66 (4.0%) | 0.083 |
| Age at first AOM episode (days), mean (sd) |  | 592 (379.5) | 357.3 (210.7) | 0.764 |
| Infant healthcare utilization |  |  |  |  |
|  | Well child visits, mean (sd) | 11 (2.1) | 11.5 (1.8) | 0.258 |
|  | Other outpatient/telehealth encounters, mean (sd) | 10.8 (11.1) | 17.9 (11.5) | 0.627 |
|  | Any ED visit, n (%) | 2,743 (55.6%) | 856 (52.4%) | 0.065 |
|  | Any hospitalization, n (%) | 463 (9.4%) | 132 (8.1%) | 0.046 |
| Comorbidities |  |  |  |  |
|  | Atopy, n (%) | 1,343 (27.2%) | 322 (19.7%) | 0.178 |
|  | GERD, n (%) | 539 (10.9%) | 220 (13.5%) | 0.078 |
| # of vaccine doses received |  |  |  |  |
|  | PCV, mean (sd) | 3.3 (0.8) | 2.9 (0.9) | 0.480 |
|  | Seasonal influenza, mean (sd) | 1.7 (1.4) | 1.1 (1.1) | 0.484 |
|  | Hib, mean (sd) | 2.8 (0.9) | 2.6 (0.9) | 0.242 |
| AOM: acute otitis media; SMD: standardized mean difference; NICU: neonatal intensive care unit; sd: standard deviation; ED: emergency department; GERD: gastroesophageal reflux disease; PCV: pneumococcal conjugate vaccine; Hib: Haemophilus influenzae type b vaccine |  |  |  |  |

To understand how patient characteristics changed over time, we assessed healthcare utilization and vaccine and antibiotic receipt prior to each subsequent AOM episode (**Table 2**). Children with rAOM had significantly fewer days between episodes compared to children with sporadic episodes (median: 111 days vs. 320 days, SMD=0.932, **Supplemental Table 2**). Despite this, children with rAOM had a greater number of antibiotic prescriptions between AOM episodes compared to children with sporadic episodes (mean: 2.4 vs. 1.6 prescriptions; SMD=0.395). We hypothesized that the increased number of antibiotic prescriptions among children who developed rAOM could be due to treatment failure, wherein a given AOM episode requires treatment with additional antibiotics due to worsening symptoms or a lack of improvement. We defined treatment failure as a subsequent healthcare encounter with a diagnosis of AOM and a new antibiotic prescription within 3-14 days of an initial AOM diagnosis. We found that children who developed rAOM had a treatment failure rate of 11.7%, while children who did not meet criteria for rAOM had a treatment failure rate of 5.9%. Taken together, these data suggest that there are significant differences in demographics, healthcare utilization, timing and cadence of AOM episodes, and treatment failure rates among children who developed rAOM.

**Table 2.** Episode-level characteristics associated with sporadic vs. recurrent AOM*.

|  |  |  |  |
| --- | --- | --- | --- |
| Characteristics and exposures | Sporadic AOM | Recurrent AOM | SMD |
|  | N = 9127 | N = 5650 |  |
| Number of days since prior episode, mean (sd) | 319.5 (286.5) | 111.1 (134.3) | 0.932 |
| Infant healthcare utilization before each episode |  |  |  |
| Well child visits, mean (sd) | 7.8 (2.5) | 7 (2.3) | 0.335 |
| Other outpatient/telehealth encounters, mean (sd) | 11.7 (14.2) | 12.2 (10.5) | 0.041 |
| Any ED visit, n (%) | 3,698 (40.5%) | 1,719 (30.4%) | 0.212 |
| Any hospitalization, n (%) | 977 (10.7%) | 536 (9.5%) | 0.040 |
| # of vaccine doses received |  |  |  |
| PCV, mean (sd) | 3.5 (0.8) | 3.3 (0.8) | 0.278 |
| Seasonal influenza, mean (sd) | 2.1 (1.5) | 1.7 (1.3) | 0.267 |
| Hib, mean (sd) | 3.1 (0.8) | 3 (0.9) | 0.062 |
| # of antibiotics received |  |  |  |
| Antibiotic prescriptions, mean (sd) | 1.6 (2) | 2.4 (2.5) | 0.395 |
| Antibiotic administrations, mean (sd) | 0.3 (1) | 0.3 (0.8) | 0.033 |
| *Assessed for up to 4 episodes for each child |  |  |  |
| AOM: acute otitis media; SMD: standardized mean difference; sd: standard deviation; ED: emergency department; GERD: gastroesophageal reflux disease; PCV: pneumococcal conjugate vaccine; Hib: Haemophilus influenzae type b vaccine |  |  |  |

### Performance of predictive models at the time of each AOM episode

Given that rAOM is likely multifactorial, we sought to identify patterns of features associated with rAOM development based upon data available at the time of each AOM episode. We used LASSO to train episode-specific models, which used all data available up to the point of a specific episode (i.e., first episode, second episode, etc.). The number of unique patients at each assessment timepoint, outcome rate (i.e., the number and percentage of children who developed rAOM) AUROC, AUPRC, and calibration slope for each model are shown in **Table 3**, with the model AUROC shown in **Figure 2**. As expected, we observed that the performance of the models improved with each subsequent AOM episode, as rAOM is defined by the number of AOM episodes. The episode 1 model, built with data from birth up to the time of the first documented AOM episode, had an AUROC of 0.75. Among children in the test data set, 21% met criteria for rAOM; thus, an AUPRC of 0.41 indicates that the model increased the true detection rate by two-fold over chance identification. The episode 2 model, built with data from birth up to the time of the second documented AOM episode, had substantially improved performance metrics, with an AUROC of 0.80 and an AUPRC of 0.63, nearly tripling the true detection rate for children who went on to develop rAOM at the time of their second AOM episode. Overall, these models demonstrate that features captured within EHR data can distinguish children who will go on to meet criteria for rAOM from those with only sporadic episodes as early as the time of the first AOM episode.

**Figure 2.**
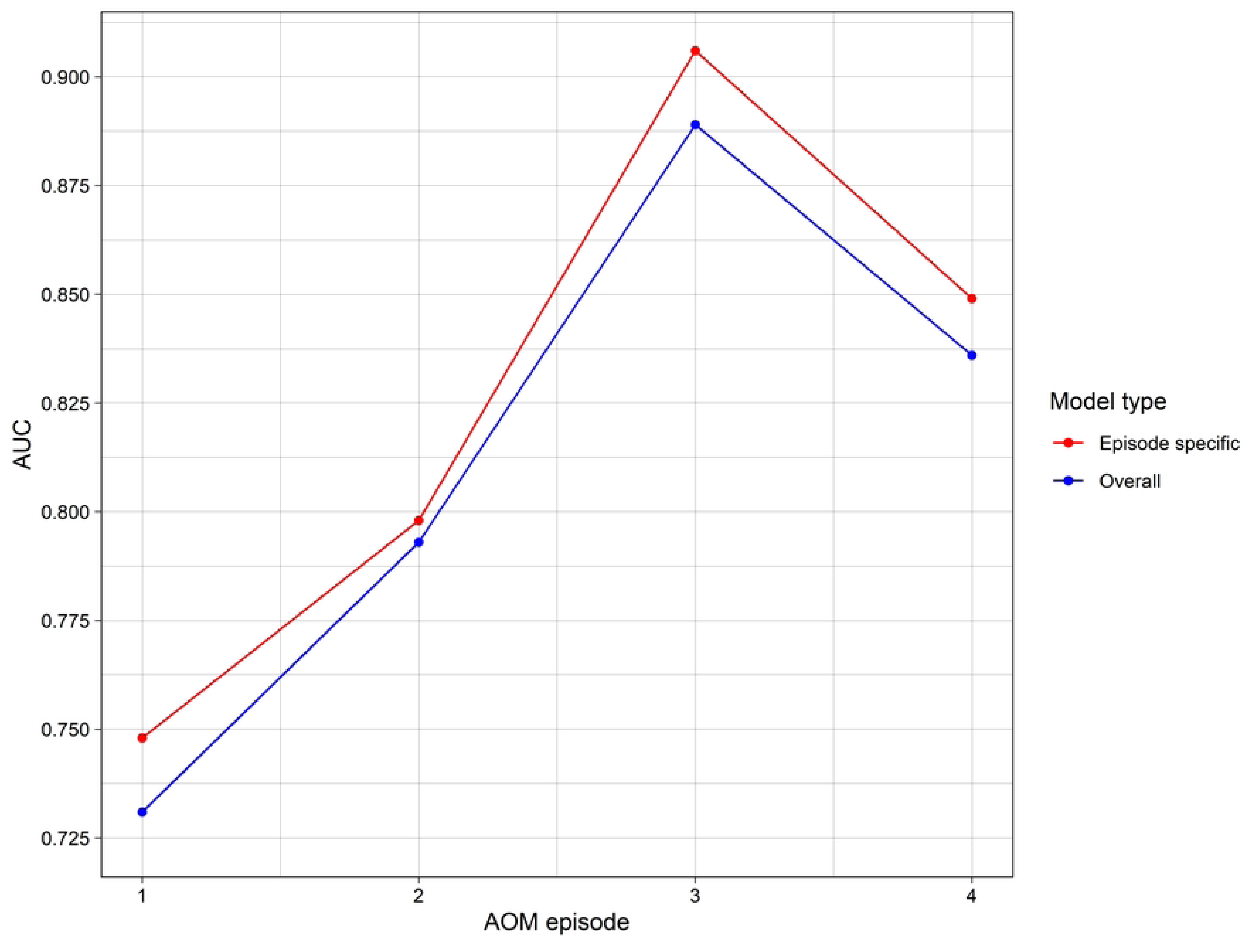
Performance of a LASSO-based model using EHR data to predict which children will meet criteria for recurrent AOM. LASSO was used to build two different models for prediction of rAOM: an overall model, which did not account for episode order, and four episode-specific models that only used data from birth up to the time of a particular episode. Area under the receiver operating curve (AUROC) for episode-specific (red) and the overall (blue) models.

**Figure 3.**
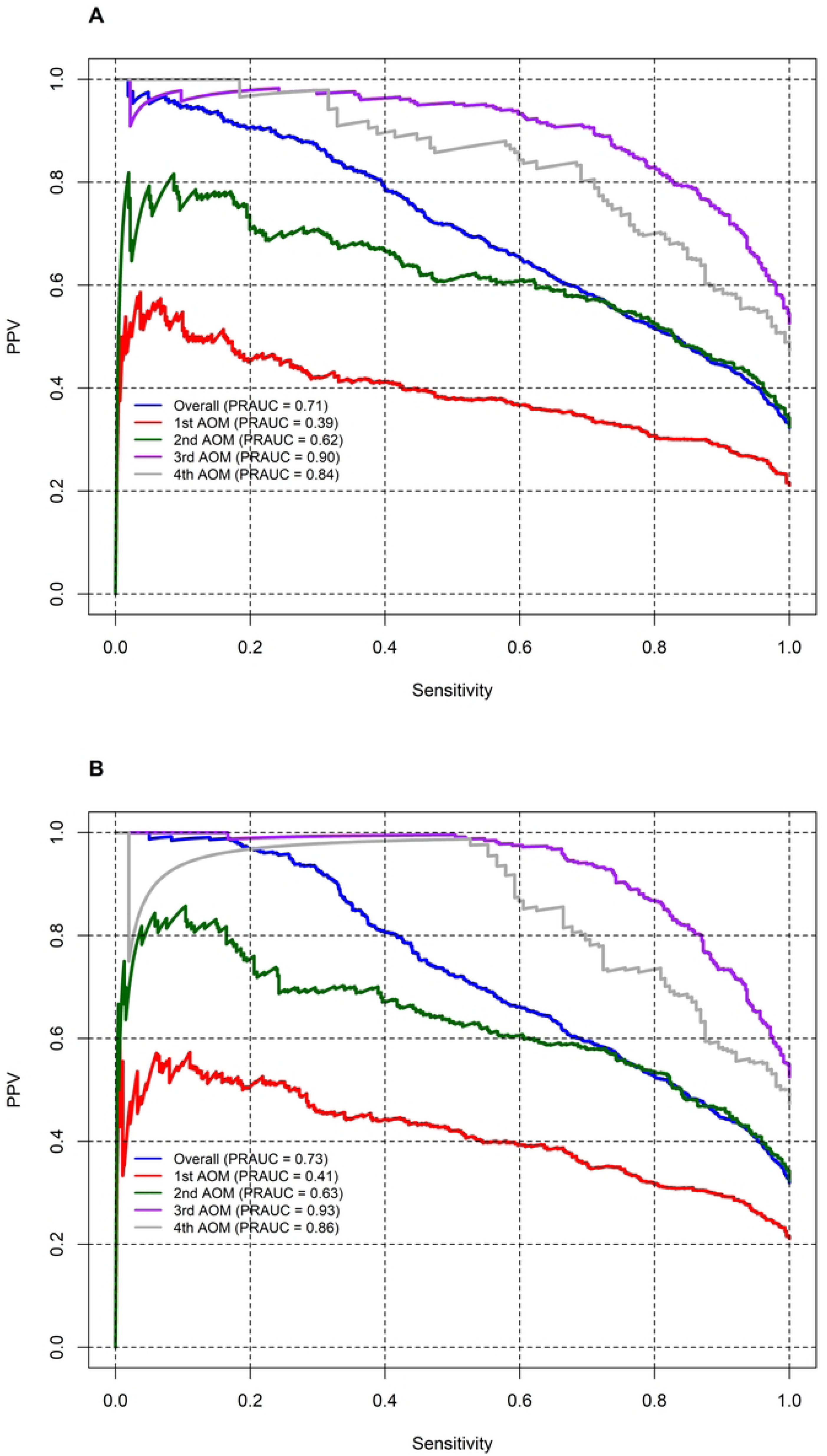
Evaluation of model performance using the Area Under the Precision-Recall Curve (AUPRC). AUPRC curves for **A**) the overall model and **B**) the episode-specific models. AUPRC estimates the average positive predictive value (i.e., the fraction of cases labeled as positive that have rAOM) and provides an understanding of how well we can identify a true positive case over random chance.

**Table 3.** AUC and calibration slopes of LASSO models.

| **Table 3. AUC and calibration slopes of LASSO models** |  |  |  |  |  |
| --- | --- | --- | --- | --- | --- |
| Model assessment timepoint | # of episodes | Outcome rate (# of children who developed rAOM) | AUC (95% CI) | Calibration slope (95% CI) | AUPRC (95% CI) |
| Episode 1 | 2189 | 462 (21.1%) | 0.75 (0.73, 0.77) | 1.07 (0.93,1.19) | 0.41 (0.37, 0.46) |
| Episode 2 | 1424 | 462 (32.4%) | 0.80 (0.77, 0.82) | 0.97 (0.89,1.06) | 0.63 (0.58, 0.68) |
| Episode 3 | 879 | 462 (52.6%) | 0.91 (0.89, 0.93) | 1.04 (0.98,1.10) | 0.93 (0.91, 0.95) |
| Episode 4 | 321 | 152 (47.4%) | 0.85 (0.80, 0.89) | 1.05 (0.91,1.18) | 0.86 (0.80, 0.90) |

### Contribution of different data features that distinguish children who will develop rAOM

To better understand the data elements that contribute to model performance and that may help distinguish children who will develop rAOM, we evaluated the LASSO coefficients for each episode-specific model and the overall model. While we cannot evaluate a causative role for these predictor variables, the coefficients provide insights into the relative importance and direction of predictor variables to the performance of each model (**Figure 4**). In the overall model, the number of AOM episodes and the number of days since a child’s first AOM episodes were the strongest predictors of rAOM. Other predictive features included child age at the time of the first AOM episode, maternal GBS-positive status at the time of delivery, the number of times children received oral antibiotics, any inpatient antibiotic administrations, and the number of outpatient healthcare visits. In our models, gastroesophageal reflux disease (GERD) diagnosis during the study period was positively associated with rAOM development, while atopy diagnoses were negatively associated.

**Figure 4.**
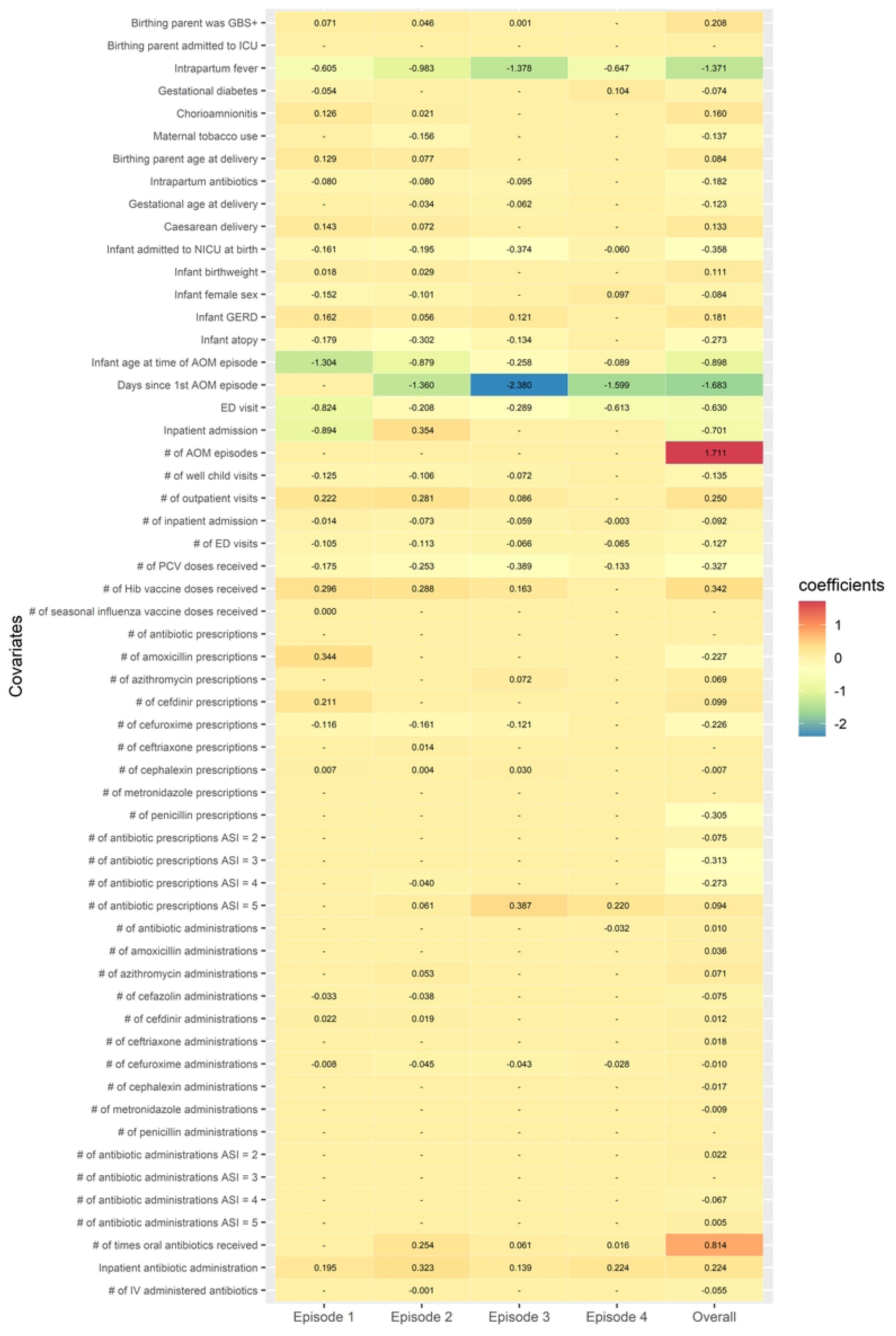
Feature coefficients of the LASSO models for prediction of rAOM at each episode and for the overall model. We evaluated the association coefficients for each model to gain an understanding of the importance of each variable to model performance. Features that were positively associated with rAOM are shown in orange-red shades, while negatively associated features are in green-blue shades. The corresponding model is listed at the bottom of the heatmap.

We additionally observed changes in the importance of predictor variables across AOM episode models. In the episode 1 model, which was built with data from birth up to the time of the first documented AOM episode, infant age, maternal intrapartum fever, and the occurrence of a prior ED visit or inpatient admission, were all negatively associated with subsequent development of rAOM, while maternal chorioamnionitis, birthing parent age at time of delivery, Caesarean delivery, number of prior outpatient visits, and number of prior amoxicillin and cefdinir prescriptions were all positively associated with subsequent rAOM development. In general, birth-associated features, such as maternal conditions, became less important with each subsequent episode. In contrast, the receipt of broad-spectrum antibiotics (the number of prescriptions with an ASI of 5) became more important in models of subsequent AOM episodes, while the importance of the number of pneumococcal conjugate vaccine doses decreased. The number of days since the first episode was strongly associated with rAOM development across all models, indicating that shorter intervals between episodes was strongly predictive. Taken together, these data indicate that the factors associated with rAOM development shift with increasing age and with subsequent episodes.

## DISCUSSION

The overarching goal of this work was to identify potential risk factors and protective factors for the development of rAOM. We used LASSO-based predictive models to identify clinical features and exposures that are associated with the subsequent development of rAOM. We built two different types of models: episode-specific models that used all available EHR data up to the point in time at which a child presented with their first, second, third, or fourth episode, and an overall model that did not account for episode order. An evaluation of factors that contributed to each model found that birth-related factors and infant age contributed to prediction performance at the time of first AOM episode, but the strength of these associations decreased in subsequent episodes. Prior antibiotic exposures, patterns of healthcare utilization, and the diagnosis of atopy and GERD also contributed to model performance. These findings indicate that clinical exposures and patient characteristics documented in the EHR may distinguish children who are at risk of developing rAOM.

Previous studies have found associations between birth mode, breastfeeding, tobacco smoke exposure, family history of AOM, presence of older siblings, and daycare attendance; however, these factors do not fully explain rAOM risk.^5^ In the present study, we identified associations with clinical exposures and sociodemographic factors that were present prior to the first AOM episode. While we could not assess the impact of breastfeeding or daycare attendance, we identified numerous other factors that were associated with the likelihood of developing rAOM, including age at first AOM episode, time between episodes, and prior antibiotic exposures.

Several previous studies have also identified associations between age at AOM onset and the development of rAOM.^41^ Bajorski and colleagues recently developed a predictor model to anticipate AOM recurrence for children with 2 or more closely spaced AOM episodes (2 or more episodes spaced no more than 6 months apart), which similarly identified age at first episode as a significant predictor, along with daycare attendance. Notably, the authors of this study used a prospective study design to stringently define AOM episodes, including tympanocentesis and microbiological confirmation. While we could not directly validate AOM diagnoses in our cohort, we similarly found that age at first episode and spacing of episodes were both significant predictors of rAOM.

Our predictive models identified associations between antibiotic treatment patterns and rAOM development. At the time of the first AOM episode, we found that a prior inpatient antibiotic administration and the number of prior amoxicillin and cefdinir prescriptions were positively associated with rAOM. Previous studies in animal models of enteric infection have found that early life antibiotic exposures are associated with altered immune development and increased infection susceptibility due to perturbation of the gut microbiome.^42,43^ Further, a study by Chapman and colleagues identified an association between antibiotic receipt in children under two years of age and decreased vaccine responses.^44^ Additionally, Toivonen and colleagues found that antibiotic treatment in early life resulted in altered nasopharyngeal microbiome developmental trajectories and asthma diagnosis later in childhood.^45^ We additionally found that children who developed rAOM had a greater number of antibiotic prescriptions between episodes and nearly double the rate of treatment failure compared to children who only had sporadic episodes. These data suggest that these episodes may be more recalcitrant and potentially involve pathogens that are less sensitive to first-line treatment approaches. Further studies will be needed to understand the underlying microbiologic features in children with rAOM and to assess how antibiotic treatment in early life influences the microbiome, pathogen susceptibility, and immune development.

This study has several strengths and limitations. Strengths include the large cohort size and inclusion of children who received consistent care within the same health system throughout the first 4 years of life. Limitations include the use of only a single center and the inability to stringently define AOM episodes, relying only on documentation within the EHR. Further, our finding that rAOM prevalence differs by race/ethnicity may reflect differences in healthcare access and utilization or in differences in diagnostics and treatment associated with race/ethnicity.^46,47^ Thus, the applicability of our findings may be limited in non-white populations and/or populations with limited healthcare access.

In conclusion, we used predictive modeling approaches to identify features that are associated with the development of rAOM in early childhood. These findings suggest that additional risk factors, such as early life antibiotic exposures and GERD may directly contribute to rAOM risk. Additionally, our results suggest that it may be possible to predict which children are likely to go on to develop rAOM as early as their first AOM episode. Such models could be deployed within EHR systems to identify children who would benefit from early evaluation by an otolaryngologist and audiologist. Early identification could enhance the impact of tympanostomy tube placement by enabling early referral of children while they are at greatest risk of AOM-related hearing loss, reducing the burden of rAOM for young children and their families.

## Data Availability

All relevant data are within the manuscript and its Supporting Information files.

## Abbreviations

AOM: acute otitis media
rAOM: recurrent acute otitis media
AUC: area under the curve
AUPRC: area under the precision-recall curve

## Conflict of Interest Disclosures (includes financial disclosures)

SAG has received paid research travel from MedEl. CWW. has intellectual property and equity with Biomeme Diagnostics, is a consultant for Emit Bio, and serves on data safety and monitoring boards for Bavarian Nordic, Meiji Seika Pharma Co., and Jannsen Pharmaceuticals. MSK is a consultant for Merck & Co, Inc. MJS is a consultant for and has received research support from Merck & Co, Inc., and Pfizer, Inc. The other authors have no conflicts of interest to disclose.

## Role of Funder/Sponsor (if any)

The funders had no role in the design and conduct of the study.

## Clinical Trial Registration (if any)

Not applicable

## Contributors Statement

Dr Jillian Hurst conceptualized and designed the study, drafted the initial manuscript, and critically reviewed and revised the manuscript.

Congwen Zhao collected the data, conducted the statistical analyses, and critically reviewed and revised the manuscript.

Drs Eileen Raynor, Janet Lee, Sarah Gitomer, Christopher Woods, Matthew Kelly, and Michael Smith, critically reviewed and revised the manuscript for important intellectual content.

Dr Ben Goldstein designed the study and critically reviewed and revised the manuscript.

All authors approved the final manuscript as submitted and agree to be accountable for all aspects of the work.

